# Epidemiology of Cannabis Use Among Middle-Aged and Older Adults in the United States

**DOI:** 10.1101/2024.10.11.24315329

**Authors:** Ofir Livne, Malka Stohl, Jodi Gilman, Terry E. Goldberg, MM Wall, Deborah S. Hasin

## Abstract

**Importance:** Studies report disproportionate increases in cannabis use among middle-aged (50-64) and older (65+) U.S. adults, groups particularly vulnerable to its adverse effects. However, national data on attitudes towards cannabis use and legalization, as well as prevalences of key cannabis-related behaviors are lacking for these groups.

**Objective:** To present national prevalences of past-year any cannabis use, medical use, consumption methods, and other important use-related behaviors, and attitudes toward use, as well as associations of such behaviors and attitudes to cannabis use among U.S. adults ages 50 and older.

**Design:** Cross-sectional data from the Health and Retirement Study (HRS) were used to calculate weighted prevalences for all cannabis measures by primary age groups (50-64, ≥65) as well as two specified older age groups (65-74, ≥75) and sex. Associations between sociodemographics and cannabis use were evaluated using multivariable logistic regression.

**Main Outcomes and Measures:** Self-reported past-year cannabis use (outcome), consumption methods, medical use and health conditions for which cannabis was used, prescriptions by healthcare providers, current and past perception of acceptability, perceptions of risk, and attitudes toward legalization (exposures). Covariates included sex, race/ethnicity, household income, and employment status.

**Setting and Participants:** HRS participants who completed the 2018 cannabis use experimental module (n=1,324).

**Results:** Past year cannabis use was reported by 18.5% (SE=2.17) and 5.9% (SE=1.19) of middle-age and older adults, respectively in the U.S. In both groups, a majority of individuals consumed cannabis exclusively by smoking. Approximately 25% of middle-aged adults and 20% of older adults used cannabis for medical purposes, with ∼20% in both groups receiving a prescription or recommendation for medical use from a healthcare provider. Over 75% of individuals in both age groups viewed medical use of cannabis as acceptable, and older adults were more likely to view cannabis as a gateway drug and to support restrictions of cannabis laws.

**Conclusions and Relevance:** Cannabis use among both middle-aged and older U.S. adults is higher than previously reported in state- and national-level studies, with many engaging in cannabis behaviors associated with increased harm. Greater public health and clinical efforts are needed for tailored prevention and intervention strategies.

## Introduction

Adults age 50 and older are more vulnerable than younger adults to numerous adverse effects of cannabis use,^1^ e.g., falls,^2^ motor vehicle crashes,^3^ injuries, emergency department visits,^4^ and declines in physical and cognitive health^5–7^. Since 2006, cannabis use has increased disproportionately among middle-aged (ages 50-64) and older (ages 65+) U.S. adults,^8–12^ with past-year use in adults 50 years and older increasing more than 3-fold.^12,13^ The vulnerability to adverse effects of cannabis use in these age groups suggests the need for greater knowledge about cannabis use behaviors to effectively address important individual and public health challenges.

Several factors may have contributed to the increased prevalence of cannabis use among adults 50 years and older. One is the aging of the Baby Boomer cohort, who had higher adolescent rates of drug use and more tolerant attitudes toward cannabis than previous cohorts.^14–19^ Additional potential contributors include decreasing perceptions of cannabis as risky^20^ and growing beliefs in its health benefits^21,22^ despite limited efficacy.^1,23^ U.S. cannabis legalization and the creation of commercial markets may also contribute to this trend, with 24 states having legalized cannabis use for recreational purposes and 38 states having legalized it for medical purposes; ∼50% of middle-aged and older adults support medical cannabis laws (MCL) and recreational cannabis laws (RCL).^24^ As more states enact cannabis laws (CL) and the U.S. population ages,^25^ the number of middle-aged and older adults consuming cannabis is expected to increase further.^12,26^ However, existing epidemiological studies of these age groups have not addressed key cannabis behaviors and related measures such as cannabis use for medical purposes (‘medical cannabis use’).^27^

Extant knowledge about cannabis use in middle-aged and older adults often relies on data from small, non-representative samples^28–31^ or from the National Survey on Drug Use and Health (NSDUH),^8–10,32^ which surveys basic measures of cannabis use but fails to capture many potential predictors of harms such as methods of consumption and motivations for use. Additionally, previous studies assessing sociodemographic characteristics of adults aged 50 years and older who use cannabis are either outdated or are based on small and non-representative samples.^33^ Consequently, many questions about cannabis use among middle-aged and older adults remain unanswered. These include: 1. What are the preferred cannabis products and consumption methods in these age groups? ^28–31^ 2. What are attitudes toward medical cannabis use, the prevalence of medical use, and the specific health conditions for which medical cannabis is being used? Also, to what extent was medical cannabis use prescribed or recommended by a healthcare provider? These questions are particularly important to answer given the risk of interactions between medication and cannabis,^34^ especially among older populations who frequently engage in polypharmacy.^35^ 3. What are key sociodemographic characteristics of middle-aged and older adults who are at increased risk of cannabis use. Considering the broader societal acceptance of cannabis that may differentially impact sociodemographic subgroups over time,^10^ research aimed at addressing this question is crucial for developing targeted approaches for screening, risk assessment, and intervention strategies.

The Health and Retirement Study (HRS),^36^ supported by the National Institute on Aging and the Social Security Administration, conducts national surveys of U.S. adults aged 50 years and over. In 2018, HRS conducted a comprehensive assessment of cannabis use behaviors, offering extensive nationally representative data on cannabis use among middle-aged and older adults in the U.S. This, together with careful sampling of these specific age groups and assessments of key sociodemographic factors relevant to middle-aged and older adults, such as employment and retirement status, and receipt of social security benefits, also make HRS a uniquely informative source of epidemiological data on cannabis use in these understudied age groups. A recent HRS brief report^37^ noted associations of cannabis use measures with health conditions, opioid use and service utilization in adults 50 years and older. However, while a useful start on addressing the epidemiology of cannabis use in this age group, the study did not report on key cannabis use measures such as methods of consumption or medical use, nor did it examine associations with age-specific sociodemographic measures, such as retirement status. Such information is critical to provide a better understanding of the cannabis landscape as it pertains to middle-aged and older adults. Therefore, using 2018 HRS data, we investigated (1) national prevalences of key cannabis behaviors never reported for adults ages 50 and older, including methods of consumption, medical cannabis use and specific conditions for which cannabis was used, healthcare provider prescription or recommendation for medical cannabis use, and perceptions and attitudes toward cannabis use; (2) whether these cannabis behaviors differed by age groups that had been combined in previous studies, and by sex; and (3) whether certain key sociodemographic characteristics are associated with increased risk of past-year cannabis use.

## Methods

### Data Source

The HRS is a longitudinal, biannual nationally representative health survey of ∼20,000 U.S. adults ages 50+ years. It employs a multi-stage area probability design with geographical stratification and clustering, and oversamples specific demographic groups,^38^ successfully recruiting and retaining minority participants.^39^ Core interviews are conducted in-person or by phone. The HRS refreshes its sample with younger cohorts every several years, with the most recent refresh in 2016.The HRS includes occasional specialized experimental modules on specific topics, administered to randomly selected participant subsamples,^40^ including one on *attitudes toward and use of Cannabis* in 2018. All study participants provided written informed consent, and protocols were approved by the University of Michigan Institutional Review Board. In the current study, we utilized data from de-identified public files, creating an analytic sample comprised of core survey respondents who participated in the 2018 experimental module. Response rates for the core survey were 74%,^41^ and sampling weights were applied to account for differential probability of selection and non-response.^38^

### Measures

#### Cannabis behaviors

Participants were assessed for past-year and lifetime cannabis use. Nonresponse on these items was imputed to ‘no use’. Among those who reported lifetime cannabis use, several variables were assessed: (1) Method of consumption (smoking, other than smoking); (4) Cannabis use for the treatment of health problems (‘medical cannabis use’); and (5) Healthcare provider recommendation for medical cannabis (‘healthcare provider involvement’). Participants reporting medical cannabis use were asked about health conditions/symptoms for which they used cannabis; these were categorized into 4 groups: emotional and psychological conditions, neurological and sensory conditions, physical conditions (including cancers/tumors; skin conditions; musculoskeletal system/connective tissue conditions; cardiovascular/blood conditions; allergies; sinusitis/tonsillitis; endocrine conditions, metabolic and nutritional conditions; gastrointestinal conditions; reproductive system conditions), and miscellaneous and other health conditions.

#### Attitudes toward cannabis use

Participants of the 2018 cannabis module, regardless of whether they reported lifetime or past year cannabis use, were asked binary questions about whether they: (1) currently believe that cannabis use for medical reasons is acceptable; (2) believed at age 18 that cannabis use for medical reasons is acceptable; (3) believe cannabis use leads to use of harder drugs (i.e., “gateway drug”); and (3) believe enforcement of cannabis laws should be restricted.

#### Sociodemographic characteristics

These included: sex (male, female); age (2-level variable representing middle-aged and older adults [51-64 years, ≥65 years] and more detailed 3-level variable [51-64 years, 65-74 years, ≥75 years]); race (White, Black or African American, other); ethnicity (Hispanic, non-Hispanic), marital status (married, annulled/separated/divorced, widowed, never married); veteran status (veteran, non-veteran); educational level (less than high school; high school, some college or higher); employment and retirement status (employed, unemployed, retired); annual household income ($0-$19,999, $20,000-$34,999, $35,000-$59,999, ≥$60,000); poverty level (below, above), defined according to yearly U.S. Census poverty thresholds; and receipt of social security benefits.

### Statistical Analysis

Weighted cross-tabulations were used to produce estimated means, prevalences, and standard errors for all cannabis behavior measures within two primary age groups (50-64, ≥65), as well as in two specified older age groups (65-74, ≥75), overall and by sex. Differences in means and prevalences between age groups and between sexes within each age group were evaluated using Wald chi-square tests. Associations between sociodemographic variables and past-year cannabis use were estimated using odds ratios (ORs) obtained from multivariable logistic regressions that controlled for sex, continuous age, race, and educational level. Analyses were performed using SAS 9.4 and SUDAAN version 11.0 to account for the HRS complex sample design.^42^

## Results

### Sample characteristics

Among HRS participants of the 2018 experimental module on cannabis use behaviors, 54.6% (SE=2.07) were ages 51-64 and 45.4% (SE=2.07) were ages 65+. e-Table 1 presents the sociodemographic characteristics of the analytic sample, overall and by age group

### Cannabis measures by age and sex (Table 1)

**Table 1.**
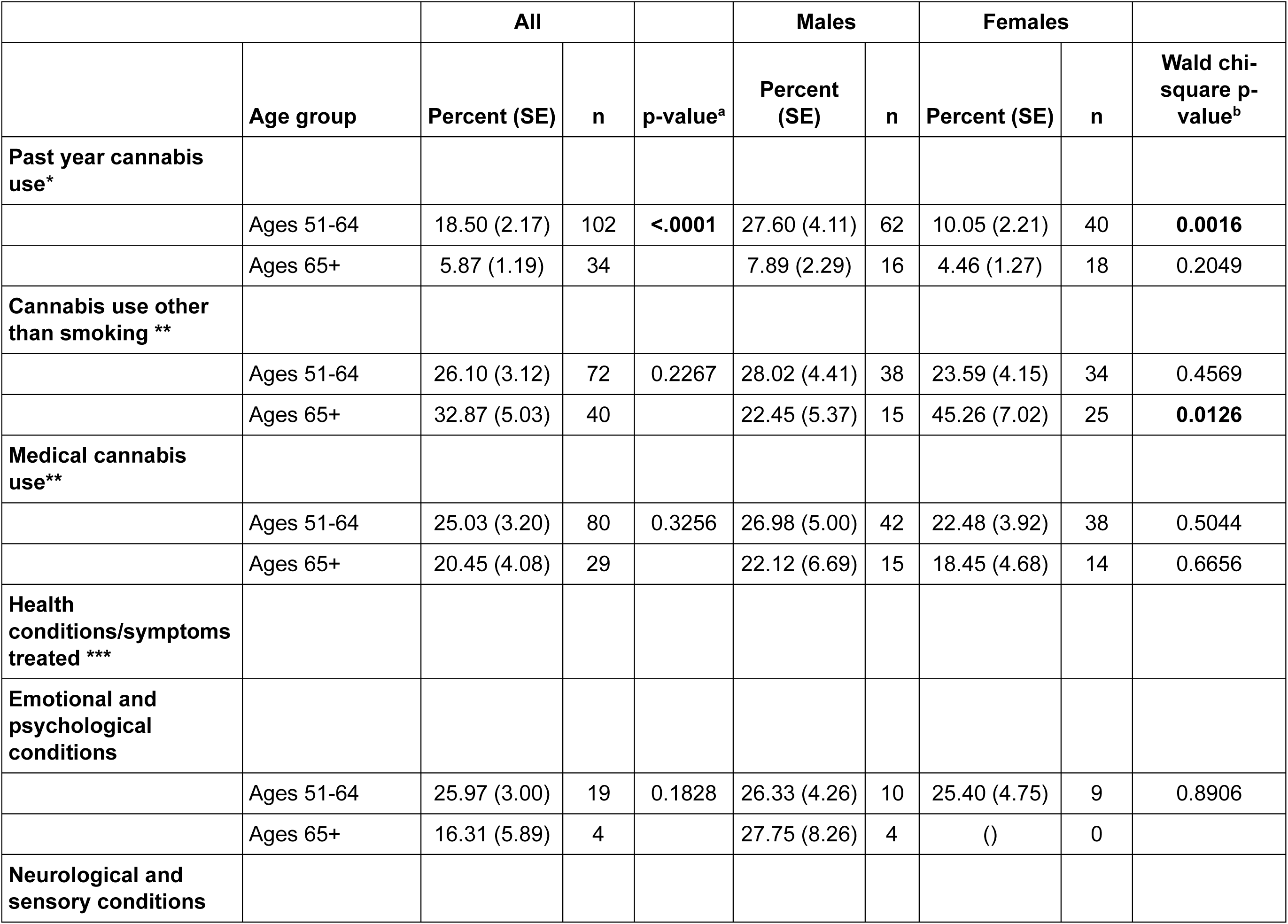
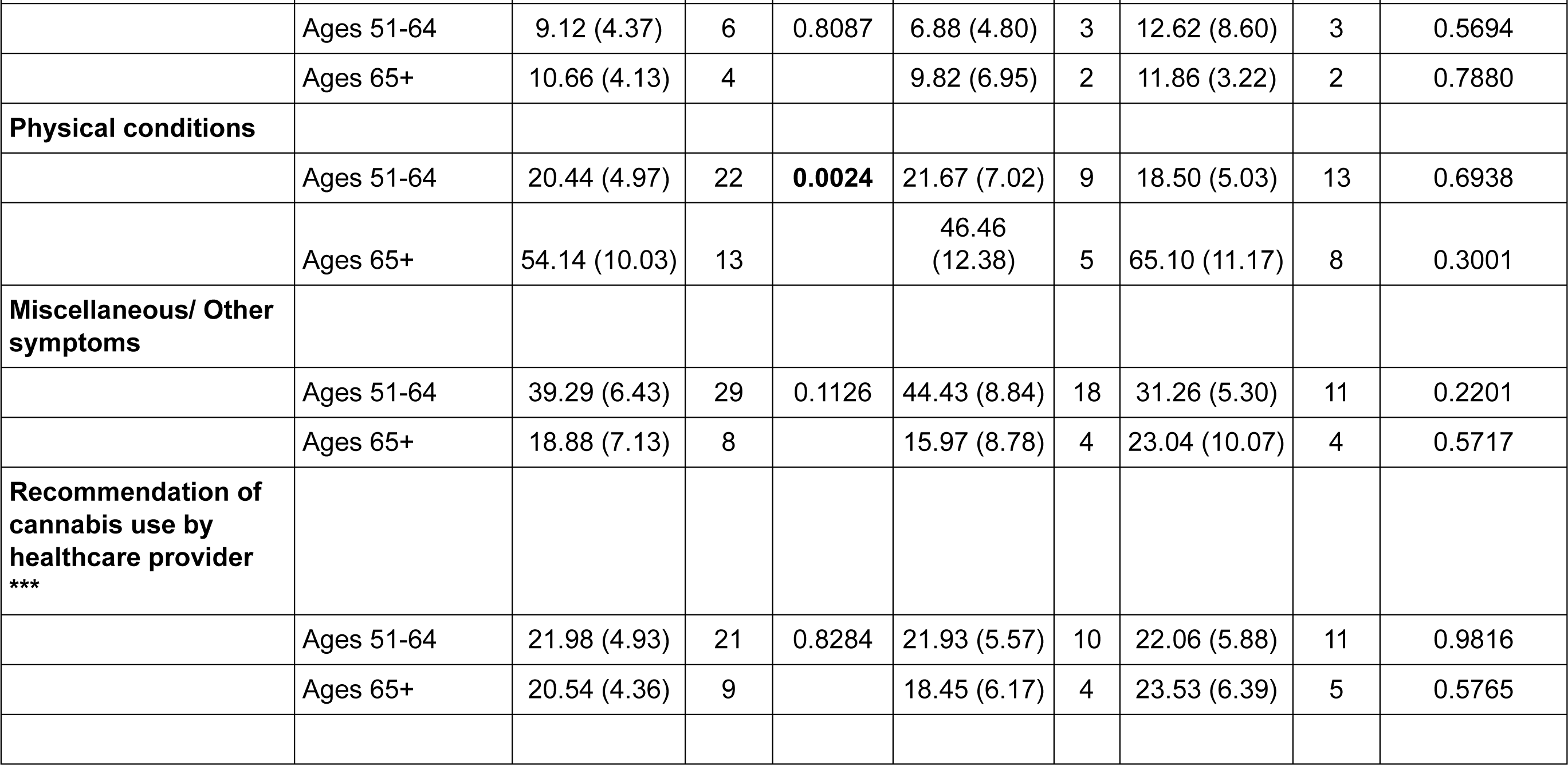
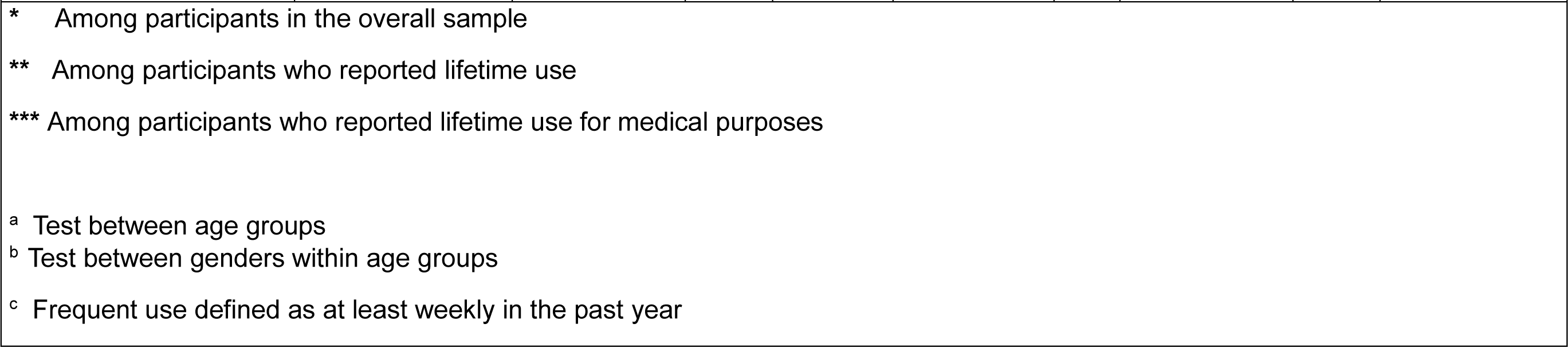
Prevalence of cannabis use measures by age (51-64, 65+) and by sex.

The prevalence of past-year cannabis use was 18.50% (SE=2.17) among middle-aged adults and 5.87% (SE=1.19) among older adults (p<.0001). Among adults age 65+, prevalences were higher in those age 65-74 (8.84% [SE=2.01]) than in those 75+ (1.78% [SE=0.60]), p<.0001; e-Table 2).

Among individuals reporting lifetime cannabis use, 26.10% (SE=3.12) of middle-aged adults and 32.87% (SE=5.03) of older adults consumed cannabis through non-smoking methods (p=0.23).

Among individuals reporting lifetime cannabis use, 25.03% (SE=3.20) of middle-aged adults and 20.45% (SE=4.08) of older adults reported lifetime medical cannabis use (p=0.33). Health conditions for which medical cannabis was used appear in Table 1. Among middle-aged adults, miscellaneous and other conditions (39.29%) were the most commonly reported reasons for medical cannabis use, followed by emotional and psychological conditions (25.97%), physical conditions (20.44%), and neurological and sensory conditions (9.12%). Older adults most commonly reported physical conditions (54.14%) as their reason for medical cannabis use, followed by miscellaneous and other conditions (18.88%), emotional and psychological conditions (16.31%), and neurological and sensory conditions (10.66%). Among medical cannabis users, similar proportions of middle-aged and older adult users received a healthcare provider’s recommendation for medical cannabis (21.98% [SE=4.93] and 20.54% [SE=4.36] respectively (p=0.82).

Few sex differences in cannabis behaviors were found. Among middle-aged adults, the prevalence of past-year cannabis use was nearly three times higher in males than in females (males: 27.60% [SE=4.11]; females: 10.05% [SE=2.21], p<.01). Among older adults, the prevalence of cannabis consumption through non-smoking methods was twice as high in females compared to males (males: 22.45% [SE=5.37]; females: 45.26% [SE=7.02], P<.05).

### Attitudes toward cannabis by age and sex (Table 2)

**Table 2.** Attitudes towards cannabis use by age (51-64, 65+) and by sex.

|  |  | All |  |  | Males |  | Females |  |  |
| --- | --- | --- | --- | --- | --- | --- | --- | --- | --- |
|  | Age group | Percent (SE) | n | p-value<sup>a</sup> | Percent (SE) | n | Percent (SE) | n | Wald chi-square p-value<sup>b</sup> |
| **Current belief that medical cannabis use is acceptable** |  |  |  |  |  |  |  |  |  |
|  | Ages 51-64 | 89.12 (1.76) | 511 | **<.0001** | 91.69 (1.92) | 230 | 86.70 (2.66) | 281 | 0.1185 |
|  | Ages 65+ | 77.19 (2.44) | 515 |  | 75.98 (3.82) | 208 | 78.04 (2.57) | 307 | 0.6114 |
| **Past (at age 18) beliefs that medical cannabis use is acceptable** |  |  |  |  |  |  |  |  |  |
|  | Ages 51-64 | 39.74 (2.68) | 216 | **<.0001** | 44.04 (3.83) | 111 | 35.72 (3.42) | 105 | 0.0967 |
|  | Ages 65+ | 13.84 (1.86) | 79 |  | 19.62 (3.21) | 46 | 9.67 (2.23) | 33 | **0.0191** |
| **Current belief that cannabis is a gateway drug** |  |  |  |  |  |  |  |  |  |
|  | Ages 51-64 | 41.59 (3.00) | 293 | **<.0001** | 39.81 (4.51) | 120 | 43.29 (4.00) | 173 | 0.5640 |
|  | Ages 65+ | 71.18 (2.12) | 490 |  | 67.50 (3.34) | 192 | 73.76 (2.75) | 298 | 0.1706 |
| **Current belief in need to restrict enforcement of cannabis laws** |  |  |  |  |  |  |  |  |  |
|  | Ages 51-64 | 37.62 (2.99) | 273 | **<.0001** | 34.26 (4.04) | 111 | 40.78 (3.87) | 162 | 0.2142 |
|  | Ages 65+ | 55.75 (2.89) | 393 |  | 53.09 (4.36) | 156 | 57.65 (3.66) | 237 | 0.4176 |
|  | Age group | Percent (SE) | n | p-value<sup>a</sup> | Percent (SE) | n | Percent (SE) | n | Wald chi-square p-value<sup>b</sup> |
| <sup>*</sup> All questions on attitudes towards cannabis use were assessed in the overall sample<br><sup>a</sup> Test between age groups<br><sup>b</sup> Test between genders within age groups |  |  |  |  |  |  |  |  |  |

In 2018, 89.12% (SE=1.76) of middle-aged adults and 77.19% (SE=2.44) of older adults viewed medical cannabis use as acceptable (p<.0001). At age 18, 39.74% (SE=2.68) of middle-aged adults and 13.84% (SE=1.86) of older adults viewed medical cannabis use as acceptable (p<.0001). Older adults were significantly more likely than middle-aged adults to view cannabis as a gateway drug (51-64: 41.59% [SE=3.00]; 65+: 71.18% [SE=2.12], P<.0001). e-Table 3 shows additional information about attitudes toward cannabis among those ages 65-74 and 75+. With few exceptions, no significant sex differences were observed in attitudes toward cannabis in either age group.

### Sociodemographic variables (Table 3)

**Table 3.** Associations between sociodemographic variables and past-year cannabis use among adults ages 50-64 and 65+.

|  | 50-64 | 65+ |
| --- | --- | --- |
|  | OR (95% CI)* | OR (95% CI)* |
| **Gender** |  |  |
| Females vs. Males | **0.30 (0.15, 0.61)** | 0.60 (0.24, 1.48) |
| **Race** |  |  |
| Black vs. White | 0.65 (0.34, 1.25) | 0.65 (0.22, 1.91) |
| Other race vs. White | 0.54 (0.22, 1.33) | (, ) |
| **Ethnicity** |  |  |
| Hispanic vs. Non-Hispanic | 0.67 (0.24, 1.83) | 0.12 (0.01, 1.21) |
| **Marital Status** |  |  |
| Annulled/separated/divorced vs. Married | 1.23 (0.61, 2.48) | 2.16 (0.56, 8.32) |
| Widowed vs. Married | 1.80 (0.37, 8.77) | 2.26 (0.57, 8.97) |
| Never Married vs. Married | 1.43 (0.56, 3.69) | 5.49 (0.97, 31.12) |
| **Veteran Status** |  |  |
| Yes vs. No | 0.88 (0.33, 2.31) | 0.24 (0.05, 1.16) |

|  | **50-64** | **65+** |
| --- | --- | --- |
|  | **OR (95% CI)\*** | **OR (95% CI)\*** |
| **Educational level** |  |  |
| Highschool vs. less than high school | 0.58 (0.21, 1.57) | 5.05 (0.53, 47.82) |
| Some college or higher vs. less than high school | 0.79 (0.30, 2.06) | 7.51 (0.88, 63.85) |
| **Employment and Retirement Status** |  |  |
| Unemployed vs. Employed | 1.21 (0.60, 2.45) | 0.64 (0.15, 2.84) |
| Retired vs. Employed | 1.20 (0.49, 2.94) | 1.07 (0.41, 2.83) |
| **Annual household income** |  |  |
| \$0-19,999 vs, \$60,000 or greater | 1.23 (0.57, 2.63) | 1.06 (0.28, 4.03) |
| \$20,000-34,999 vs, \$60,000 or greater | 1.46 (0.49, 4.37) | 1.35 (0.35, 5.24) |
| \$35,000-59,999 vs, \$60,000 or greater | 0.95 (0.37, 2.44) | 0.39 (0.11, 1.45) |
| **Poverty Level** |  |  |
| Below poverty level vs. Above | 0.95 (0.36, 2.49) | 0.41 (0.09, 1.89) |
| **Social Security Benefits** |  |  |
| Yes vs. No | 1.78 (0.88, 3.60) | 1.19 (0.14, 10.44) |
| OR = Odds Ratio; CI = Confidence Interval.<br>Note: all ORs are adjusted for gender, age, education and race. |  |  |

There were no differences by sociodemographic characteristics in either age group except for sex differences in past-year cannabis use among middle-aged adults.

## Discussion

In 2018, 18.5% of U.S. adults aged 50–64 and 5.9% of U.S. adults aged 65 and older used cannabis, representing over 11.5 million and 3 million individuals, respectively.^43^ In addition to revealing higher numbers of U.S. middle-aged and older adults with past-year cannabis than previously reported in national surveys, this is the first nationally-representative epidemiological study to report the prevalences of key cannabis use behaviors in these age groups, including medical cannabis use for specific health conditions healthcare provider prescriptions. This study’s findings also highlight that a significant portion of users from these vulnerable age groups are engaging in cannabis behaviors associated with increased harm, such as smoking as a primary method of consumption.

The prevalence of cannabis use among U.S. middle-aged and older adult HRS participants is markedly higher than reported by NSDUH participants from similar age groups in corresponding years.^44,45^ While both surveys employ a multistage area probability sample design, HRS oversamples adults age 65+ from minority groups and certain selected geographic areas, whereas NSDUH focuses on broad national and state-level representation. Furthermore, HRS uses clustering and stratification variables to account for the geographic clustering of its sample, which is less emphasized in NSDUH. This enhances HRS’s variance estimates for its older adult population.^46,47^ Consequently, HRS’s probability sampling method likely offers considerable accuracy in representing middle-aged and older adults nationally.

This study separated older adults into two groups, ages 65-74 and 75+, which were combined in previous NSDUH and HRS reports, potentially overlooking cannabis behaviors and sociodemographics unique to each group. Past-year cannabis use was nearly five times higher in adults 65-74 than in those 75+, xxxx. These rising rates among these older individuals warrant clinician awareness of their increased risk of harm, driven by aging-related physiological changes such as altered drug metabolism and neurotransmitter sensitivity.^48–50^

In 2018, middle-aged males were nearly three times more likely to use cannabis than females, aligning with other national- and state-level reports. No sex differences were observed in the older adult age group, suggesting that the previously reported gender gap in cannabis use among older adults^33^ may be narrowing, possibly due to disproportionate increases in cannabis use among women,^10^ particularly for medical purposes.^51–55^ This narrowing gap, coupled with a faster progression to problematic use (i.e., “telescoping effect”)^56,57^ and a lower likelihood of seeking treatment among women,^58^ indicates a need for prevention and treatment strategies tailored to middle-aged and older adult women.

Approximately one-quarter of middle-aged and one-third of older adults reported consuming cannabis through methods other than smoking, marking the first national epidemiological report on consumption methods among these populations. This finding contrasts with data from Washington State, which indicates a broader variety of consumption methods within these populations.^55,59^ Such differences may be attributable to easier access to diverse products in states that were early to legislate MCL and RCL, such as Washington. The high rates of cannabis consumption exclusively by smoking among middle-aged and older adults—despite smoking not being the recommended method for medical use underscore their resistance to changing established habits and their continued exposure to smoking-related harms. Notably, our findings suggest that compared to older adult males, older females may be more responsive to public health messaging about using cannabis through less harmful methods. Physicians prescribing cannabis should be aware of such age- and sex-specific cannabis use behaviors, educate patients about the potential harms of smoking, and promote safer medical alternatives for medical cannabis use.

Our findings provide valuable insights into attitudes toward medical cannabis use, its prevalence, and healthcare provider involvement among middle-aged and older U.S. adults—populations increasingly using cannabis to manage chronic conditions.^60–63^ Approximately 25% of middle-aged adults and 20% of older adults who reported lifetime cannabis use have used it for medical purposes, aligning with smaller studies.^64^ However, unlike those studies, we did not observe sex differences in medical cannabis use. Unlike NSDUH surveys, which focus on healthcare provider recommendations as a proxy for medical cannabis use, HRS directly assessed it by asking participants about cannabis use for the treatment of physical and mental health problems, including specific conditions, thus providing more accurate estimates of medical use prevalences in the U.S. Among older adults, the prevalence of medical cannabis use decreased with age, mirroring the overall age-related differences in overall (both medical and non-medical) cannabis use.

Our study reveals widespread acceptance of medical cannabis across both age groups, consistent with Pew Research and smaller studies. This acceptance has grown substantially from age 18 to the present, reflecting the increasing belief in cannabis’s therapeutic benefits, possibly influenced by industry marketing efforts, ^65,66^ and suggesting an expected rise in medical cannabis use among these age groups in the future. These high rates of medical cannabis use among middle-aged and older adults are concerning, given the associated risks, including a 20-25% prevalence of CUD among medical cannabis users, surpassing that of recreational users.^67,68^ Older adults with chronic conditions, such as chronic pain, are particularly at risk of CUD. ^69,70^ As rates of medical cannabis use rise, ongoing research is needed to monitor CUD prevalence. Notably, only ∼20% of individuals with medical cannabis use in both age groups received a recommendation from a healthcare provider for cannabis to treat health conditions, indicating widespread self-medication. This practice poses risks of incorrect dosing and dangerous drug interactions. These results may reflect the growing reliance on internet and social media for information about medical cannabis use.^71^ Stigma or legal concerns may also contribute to this. While healthcare providers generally support medical cannabis use, their knowledge of its administration and legal frameworks varies, leading to mixed views on effectiveness. ^72,73^ In the absence of evidence-based guidelines, it is vital to improve education for both patients and physician, ensure access to knowledgeable providers, and engage policymakers in developing harm prevention strategies for safer use.

Older adults are more likely than middle-aged adults to view cannabis as a gateway drug, with nearly three-quarters believing it leads to the use of harder substances. This contrasts with NSDUH data, where less than half of older adults perceive cannabis as a risky.^20^ Differences in methodology, such as NSDUH’s broader questions on perceived harm versus HRS’s focus on progression to harder substances, may account for this variation. Nonetheless, older Americans perceive cannabis as more harmful than middle-aged adults. Additionally, most older adults support restrictions on cannabis legalization, consistent with Gallup polls.^74^ Further research is needed to explore whether the risk of cannabis use among older adults varies by legalization status.

This study has several limitations. First, the 2018 experimental module on cannabis use behaviors was only administered once, preventing the assessment of trends in cannabis use behaviors over time. Second, all data are self-reported, which may introduce recall and social desirability biases. third, HRS did not assess other important measures of cannabis use, such as amount of use and product type (e.g., cannabidiol), or outcomes such as CUD. Additionally, medical cannabis use for chronic pain, an increasingly common reason for use, was not specifically assessed in HRS, limiting insights into its prevalence among these age groups. However, HRS has key strengths^40^ that mitigate these limitations, including a large, nationally representative sample of U.S. adults 50 years and older, allowing for accurate estimates of cannabis use in understudied age groups. The study also employs oversampling and targeted recruitment, resulting in response rates for minorities that are roughly equivalent to those of White individuals, and a high overall panel response rate. Furthermore, it includes a significantly broader range of cannabis measures than other national studies.

## Conclusion

Amid growing belief in the health benefits of cannabis, diminishing risk perceptions, and evolving legislation, middle-aged and older adults emerge as distinct groups warranting special attention due to their growing use of cannabis and engagement in cannabis behaviors that are associated with increased harm. Epidemiological studies such as the current study are vital to understanding the diverse profiles of middle-aged and older adult cannabis users, paving the way for targeted prevention, treatment, and policy initiatives tailored to these rapidly growing and vulnerable populations.

## Data Availability

All data produced are available online at:
https://hrsdata.isr.umich.edu/data-products/hrs-2018-core-module-4-attitude-toward-and-use-marijuana-cannabis-older-americans

---

**e-Table 1:**
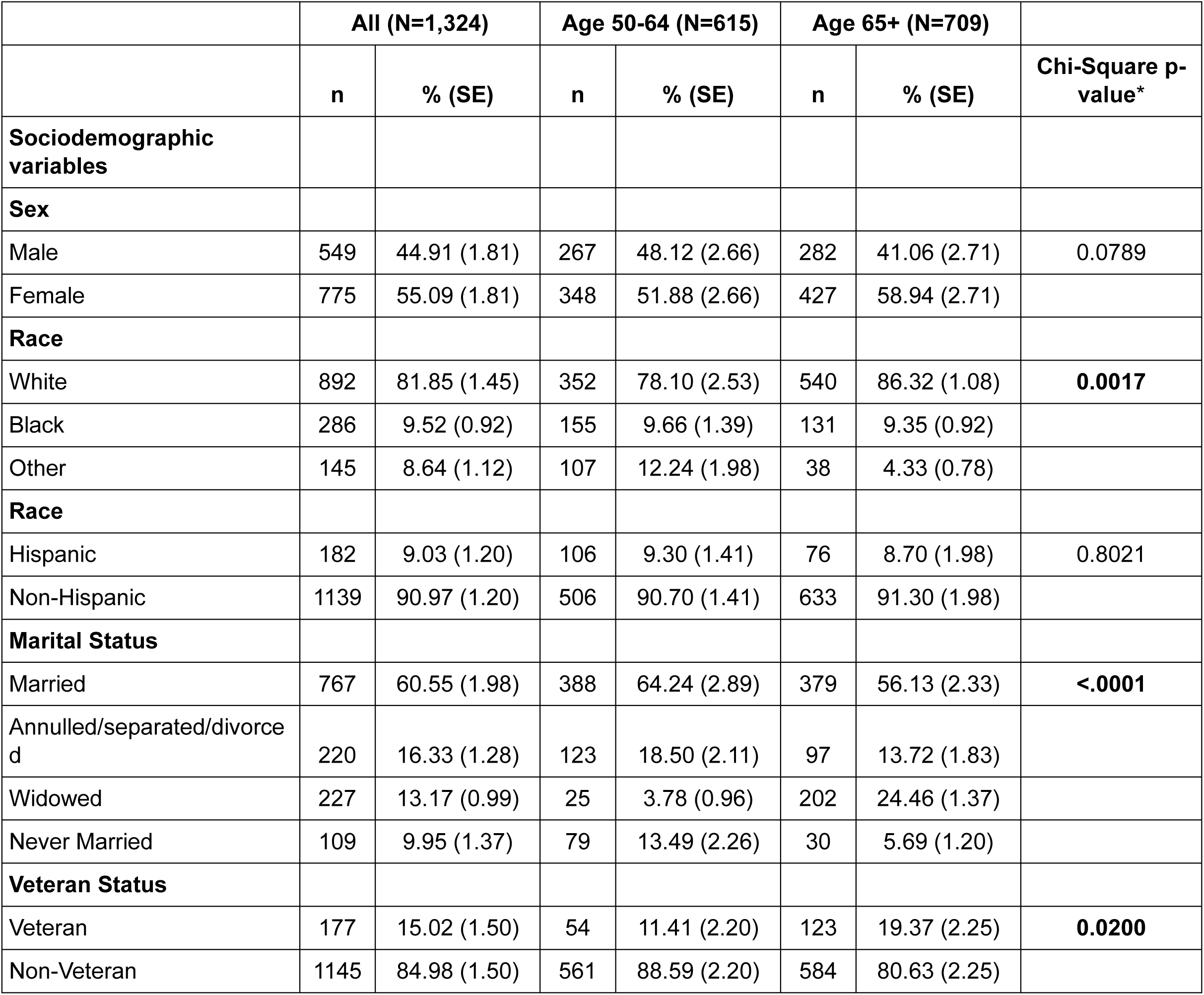
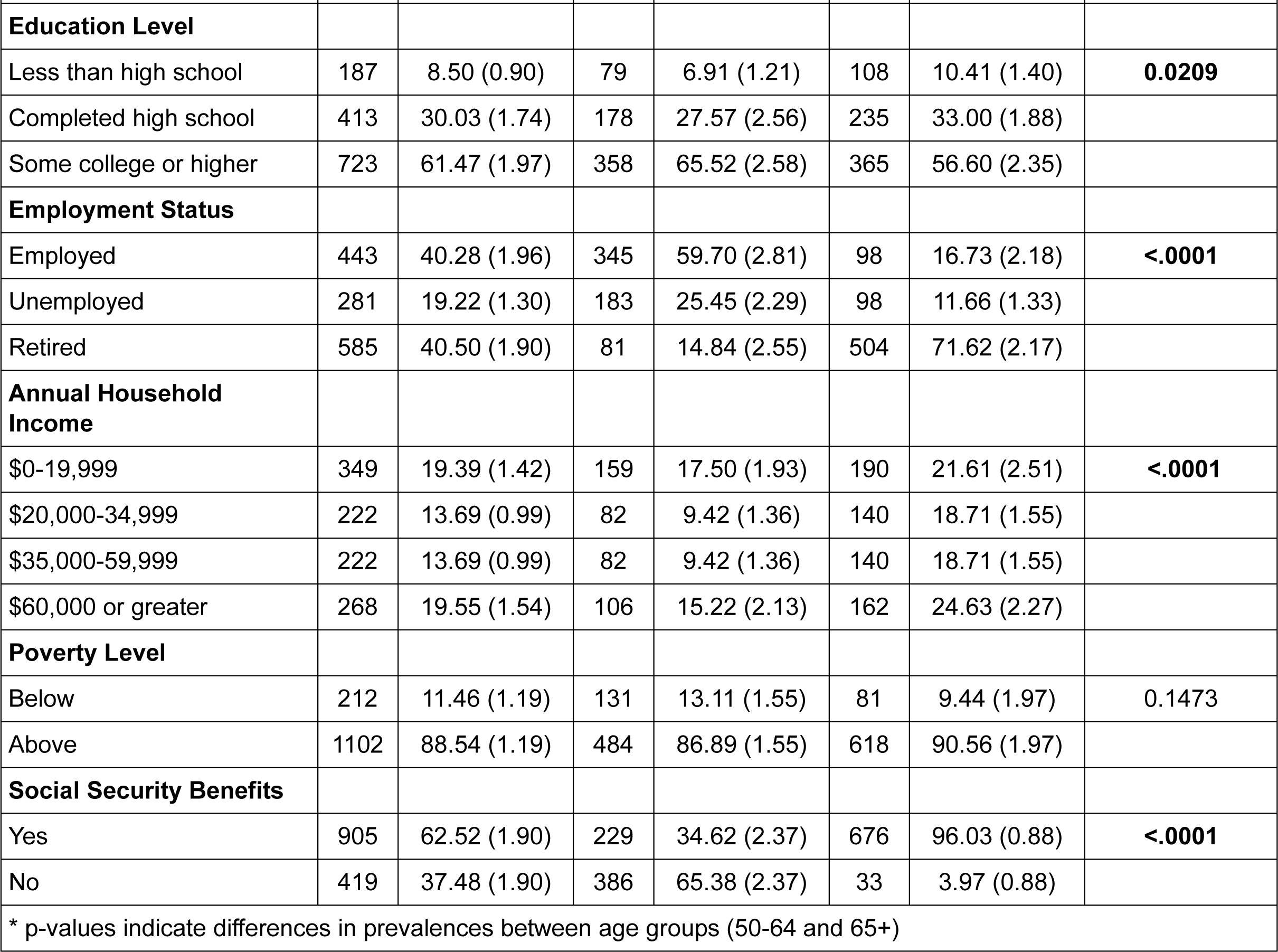
Sample descriptives by age group.

**e-Table 2.** Prevalence of cannabis use measures by age (51-64, 65-74, 75+) and by sex.

|  |  | All |  |  | Males |  | Females |  |  |
| --- | --- | --- | --- | --- | --- | --- | --- | --- | --- |
|  | Age group | Percent (SE) | n | p-value<sup>a</sup> | Percent (SE) | n | Percent (SE) | n | Wald chi-square p-value<sup>b</sup> |
| **Past year cannabis use\*** |  |  |  |  |  |  |  |  |  |
|  | Ages 51-64 | 18.50 (2.17) | 102 | **<.0001** | 27.60 (4.11) | 62 | 10.05 (2.21) | 40 | **0.0016** |
|  | Ages 65-74 | 8.84 (2.01) | 25 |  | 11.50 (3.74) | 12 | 6.81 (2.08) | 13 | 0.2722 |
|  | Ages 75+ | 1.78 (0.60) | 9 |  | 2.23 (1.11) | 4 | 1.51 (0.73) | 5 | 0.6030 |
| **Cannabis use other than smoking \*\*** |  |  |  |  |  |  |  |  |  |
|  | Ages 51-64 | 26.10 (3.12) | 72 | 0.2648 | 28.02 (4.41) | 38 | 23.59 (4.15) | 34 | 0.4569 |
|  | Ages 65-74 | 35.15 (6.00) | 29 |  | 25.36 (6.61) | 12 | 46.68 (7.92) | 17 | **0.0401** |
|  | Ages 75+ | 22.14 (6.04) | 11 |  | 9.19 (4.42) | 3 | 38.37 (12.30) | 8 | 0.0344 |
| **Medical cannabis use \*\*** |  |  |  |  |  |  |  |  |  |
|  | Ages 51-64 | 25.03 (3.20) | 80 | 0.5131 | 26.98 (5.00) | 42 | 22.48 (3.92) | 38 | 0.5044 |
|  | Ages 65-74 | 20.98 (4.89) | 21 |  | 25.06 (8.13) | 12 | 16.17 (5.55) | 9 | 0.3943 |
|  | Ages 75+ | 17.95 (6.44) | 8 |  | 8.74 (4.22) | 3 | 29.50 (14.30) | 5 | 0.1923 |
| **Health conditions/symptoms treated \*\*\*** |  |  |  |  |  |  |  |  |  |
| **Emotional and psychological conditions** |  |  |  |  |  |  |  |  |  |
|  | Age group | Percent (SE) | n | p-value<sup>a</sup> | Percent (SE) | n | Percent (SE) | n | Wald chi-square p-value<sup>b</sup> |
|  | Ages 51-64 | 25.97 (3.00) | 19 | 0.2549 | 26.33 (4.26) | 10 | 25.40 (4.75) | 9 | 0.8906 |
|  | Ages 65-74 | 17.89 (6.61) | 3 |  | 27.71 (8.28) | 3 | () | 0 |  |
|  | Ages 75+ | 7.66 (7.59) | 1 |  | 28.29 (25.29) | 1 | () | 0 |  |
| **Neurological and sensory conditions** |  |  |  |  |  |  |  |  |  |
|  | Ages 51-64 | 9.12 (4.37) | 6 | 0.3970 | 6.88 (4.80) | 3 | 12.62 (8.60) | 3 | 0.5694 |
|  | Ages 65-74 | 11.71 (4.82) | 3 |  | 10.57 (7.35) | 2 | 13.78 (4.58) | 1 | 0.6972 |
|  | Ages 75+ | 4.93 (1.82) | 1 |  | () | 0 | 6.76 (3.07) | 1 |  |
| **Physical conditions** |  |  |  |  |  |  |  |  |  |
|  | Ages 51-64 | 20.44 (4.97) | 22 | **0.0086** | 21.67 (7.02) | 9 | 18.50 (5.03) | 13 | 0.6938 |
|  | Ages 65-74 | 53.11 (10.97) | 10 |  | 50.01 (13.96) | 5 | 58.77 (15.04) | 5 | 0.6883 |
|  | Ages 75+ | 59.77 (16.54) | 3 |  | () | 0 | 81.95 (8.21) | 3 |  |
| **Miscellaneous/ Other symptoms** |  |  |  |  |  |  |  |  |  |
|  | Ages 51-64 | 39.29 (6.43) | 29 | 0.2282 | 44.43 (8.84) | 18 | 31.26 (5.30) | 11 | 0.2201 |
|  | Ages 65-74 | 17.29 (7.29) | 5 |  | 11.71 (8.38) | 2 | 27.46 (13.48) | 3 | 0.3318 |
|  | Ages 75+ | 27.64 (14.06) | 3 |  | 71.71 (25.29) | 2 | 11.29 (5.13) | 1 | 0.1614 |
| **Recommendation of cannabis use by healthcare provider**<br>*** |  |  |  |  |  |  |  |  |  |
|  | Age group | Percent (SE) | n | p-value<sup>a</sup> | Percent (SE) | n | Percent (SE) | n | Wald chi-square p-value<sup>b</sup> |
|  | Ages 51-64 | 21.98 (4.93) | 21 | 0.1750 | 21.93 (5.57) | 10 | 22.06 (5.88) | 11 | 0.9816 |
|  | Ages 65-74 | 17.21 (4.20) | 5 |  | 17.69 (5.77) | 3 | 16.33 (5.43) | 2 | 0.8666 |
|  | Ages 75+ | 38.79 (14.79) | 4 |  | 28.29 (25.29) | 1 | 42.68 (19.40) | 3 | 0.6607 |

\* Among participants in the overall sample
\*\* Among participants who reported lifetime use
\*\*\* Among participants who reported lifetime use for medical purposes
\*\*\*\* Among participants who reported past-year cannabis use
<sup>a</sup> Test between age groups
<sup>b</sup> Test between genders within age groups
<sup>c</sup> Among past-year users; frequent use defined as at least weekly in the past year

**e-Table 3.**
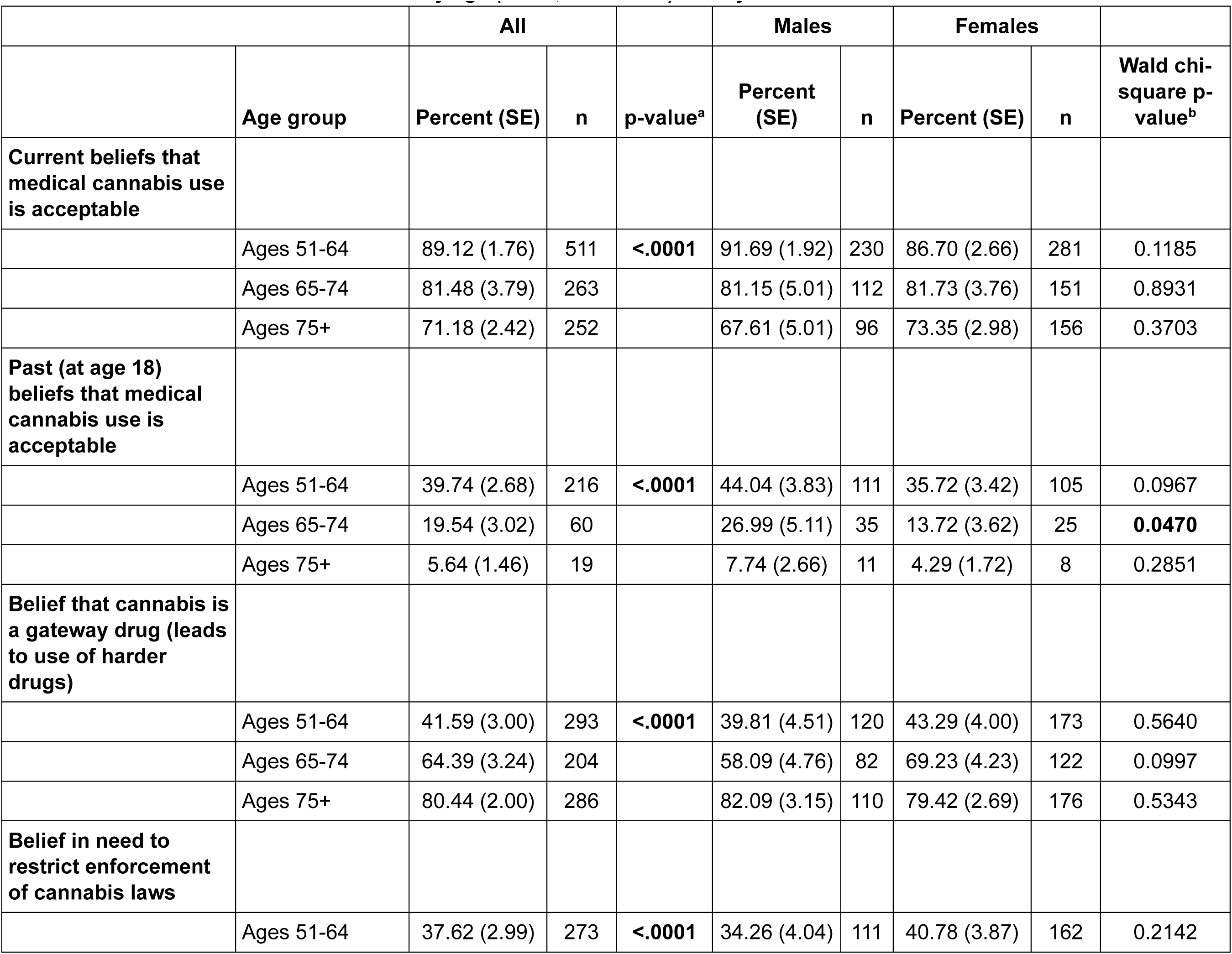
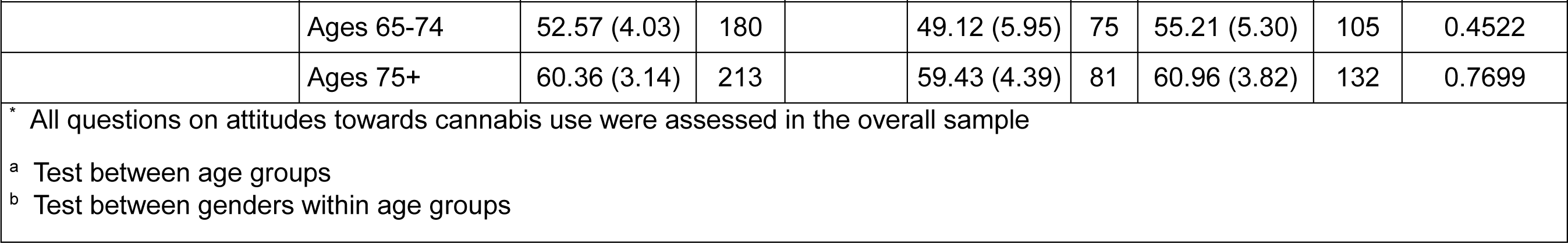
Attitudes towards cannabis use by age (51-64, 65-74. 75+) and by sex.

